# A Processed EEG based Brain Anesthetic Resistance Index Predicts Postoperative Delirium in Older Adults: A Dual Center Study

**DOI:** 10.1101/2021.01.07.21249360

**Authors:** Mary Cooter, Thomas Bunning, Sarada S. Eleswarpu, Mitchell T. Heflin, Shelley McDonald, Sandhya Lagoo-Deenadalayan, Heather Whitson, Stacie G Deiner, Miles Berger

**Author notes:** Corresponding Author: Miles Berger, MD PhD, Assistant Professor, Neuroanesthesiology Division (Duke Anesthesiology Dept.), 4317 Duke South Orange Zone, Duke University Medical Center, Durham, NC 27710. These authors contributed equally to this manuscript (Mary Cooter, Thomas Bunning). Clinical Trial Number and Registry URL: N/A. Prior presentations: N/A. Author Contributions: M.C.: This author obtained, cross-checked and analyzed data, created figures and tables, contributed written content and helped review and edit the manuscript. M.T.H.: This author helped obtain data in the Perioperative Optimization of Senior Health (POSH) clinic, assisted in analyzing data, and helped review and edit the manuscript. S.M.: This author helped obtain data in the Perioperative Optimization of Senior Health (POSH) clinic, assisted in analyzing data, and helped review and edit the manuscript. T.B.: This author reviewed data, assisted in analyzing data, and helped review and edit the manuscript. S.E.: This author reviewed data, assisted in analyzing data, and helped review and edit the manuscript. S.L.-D.: This author helped obtain data in the Perioperative Optimization of Senior Health (POSH) clinic, assisted in analyzing data, and helped review and edit the manuscript. H.W.: This author helped obtain data in the Perioperative Optimization of Senior Health (POSH) clinic, assisted in analyzing data, and helped review and edit the manuscript. M.B.: This author conceived of this project, performed study design and conception, assisted with analyzing data, and wrote the manuscript. S.D.: This author obtained data at the Mt. Sinai study site and helped review and edit the manuscript.

## Abstract

**Background:** Some older adults show exaggerated responses to drugs that act on the brain, such as increased delirium risk in response to anticholinergic drugs. The brain’s response to anesthetic drugs is often measured clinically by processed electroencephalogram (EEG) indices. Thus, we developed a processed EEG based-measure of the brain’s neurophysiologic resistance to anesthetic dose-related changes, and hypothesized that it would predict postoperative delirium.

**Methods:** We defined the Duke Anesthesia Resistance Scale (DARS) as the average BIS index divided by the quantity 2.5 minus the average age-adjusted end-tidal MAC (aaMAC) inhaled anesthetic fraction. The relationship between DARS and postoperative delirium was analyzed in derivation (Duke; N=69), validation (Mt Sinai; N=70), and combined estimation cohorts (N=139) of older surgical patients (age ≥65). In the derivation cohort, we identified a threshold relationship between DARS and for delirium and identified an optimal cut point for prediction.

**Results:** In the derivation cohort, the optimal DARS threshold for predicting delirium was 27.0. The delirium rate was 11/49 (22.5%) vs 11/20 (55.0%) and 7/57 (12.3%) vs 6/13 (46.2%) for those with DARS ≥ 27 vs those with DARS < 27 in the derivation and validation cohorts respectively. In the combined estimation cohort, multivariable analysis found a significant association of DARS <27.0 with postoperative delirium (OR=4.7; 95% CI: 1.87, 12.0; p=0.001). In the derivation cohort, the DARS had an AUC of 0.63 with sensitivity of 50%, specificity of 81%, positive predictive value of 0.55, and negative predictive value of 0.78. The DARS remained a significant predictor of delirium after accounting for opioid, midazolam, propofol, non-depolarizing neuromuscular blocker, phenylephrine and ketamine dosage, and for nitrous oxide and epidural usage.

**Conclusions:** These results suggest than an intraoperative processed EEG-based measure of lower brain anesthetic resistance (i.e. DARS <27) could be used in older surgical patients as an independent predictor of postoperative delirium risk.

## INTRODUCTION

Postoperative delirium is a common complication in older surgical patients and has been associated with increased length of stay, functional decline, and increased 1 year postoperative mortality rates.^1-3^ Recent guidelines call for^1,4^ or at least encourage^5,6^ EEG-based anesthetic management to reduce delirium rates, and intraoperative anesthetized raw EEG features such as burst suppression and alpha band power have been associated with postoperative delirium^7-10^ and preoperative cognitive impairment,^11,12^ respectively. Fritz et al demonstrated that increased anesthetic sensitivity (as indicated by EEG burst suppression at lower anesthetic dosage) is associated with increased postoperative delirium risk.^13^

However, many anesthesiologists are not familiar with raw EEG waveform analysis. As a result, many anesthesiologists currently rely on processed EEG index values to assess the neurophysiologic state of the brain in the operating room. One commonly used intraoperative processed EEG monitor is the Bispectral Index (BIS), which uses a proprietary algorithm to convert raw EEG waveforms from two frontal EEG channels into a unit-less index value between 0-100. We have recently shown that BIS index values are inaccurately high in older adults,^14^ and its use has not been shown to reduce intraoperative awareness in cases utilizing inhaled anesthetics.^15,16^

Nonetheless, the BIS is the most widely used processed EEG monitor in American operating rooms, so we reasoned that a delirium prediction tool that utilizes it would be facile to implement in the United States. We theorized that lower BIS values in response to relatively lower anesthetic doses would serve as a marker of decreased neurophysiologic resistance of the brain to the sedative/hypnotic effects of GABA-ergic anesthetic drugs, similar to the way that significant sedation in response to small amounts of alcohol is commonly viewed as a marker of lower alcohol “tolerance”. We also hypothesized that a processed EEG measure of decreased neurophysiologic resistance to GABA-ergic anesthetics would serve as a brain marker of increased postoperative delirium risk. Thus, we developed a processed EEG-based brain anesthetic resistance index based on BIS values and age-adjusted end tidal anesthetic concentrations. We then tested the hypothesis that this brain anesthetic resistance index would predict postoperative delirium risk in older surgical patients from two different institutional cohorts.

## MATERIALS AND METHODS

### Patient Population

All patients seen at the Duke perioperative optimization of senior health clinic^17^ from June 24, 2013 to September 25, 2015 were screened for inclusion into the derivation cohort for this study (N=278, see supplemental methods for additional details). This retrospective study was approved by the Duke University Medical Center IRB, which waived the informed consent requirement.

For the validation cohort, we utilized prospectively collected data from patients enrolled in an observational cohort study approved by the Mt. Sinai Medical Center IRB and registered with clinicaltrials.gov (NCT02650687). All Mt. Sinai patients underwent informed consent prior to participation, and were enrolled between November 2015 and 2018. The Mt. Sinai observational cohort study was primarily focused on postoperative cognitive dysfunction, though it also obtained postoperative delirium data. The Mount Sinai IRB waived the requirement for patients in this study to provide additional informed consent for the inclusion of their de-identified data in this manuscript. Retrospectively we extracted data collected from the Bispectral Index (BIS) monitor, intraoperative inhaled agent data and additional baseline medication information. The data was saved directly from the monitor onto a secure server.

Duke clinic and Mt Sinai patients were included in this study if they had surgery for >1 hour duration, and had end tidal anesthetic gas values and BIS index data available for more than 50% of the case minutes. To exclude total intravenous anesthetic cases, we excluded any case in which the patient received >500 mg/hr of propofol. Anesthetic case length was defined by the case start and end times documented by the anesthesia provider.

### Intraoperative Anesthetic Dosage

End-tidal anesthetic concentration (ETAC) was recorded continuously from 5 minutes after incision until 5 minutes prior to the end of surgery, in order to capture the anesthetic “plateau phase” of the case.^14^ Using a previously described method^18^ to avoid data artifacts, the end-tidal MAC fraction was recorded once per minute, and the median value over each 5-minute case epoch was obtained. The mean of these median values was then calculated to determine the overall end-tidal MAC fraction. Next, we used MAC_40_ values from our recent meta-regression analysis of age-related changes in MAC in published studies^19^ to calculate the age-adjusted end-tidal MAC fraction (aaMAC), again using the mean of median values obtained from each 5-minute case epoch.

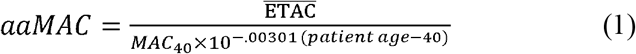

MAC-hours was defined as the product of the case duration (in hours) and the aaMAC value from equation (1).

### Bispectral Index (BIS) Value

The bispectral Index (BIS) (Covidien, Mansfield, MA) was utilized for all cases at both institutions; processed EEG values from BIS electrodes placed on the left forehead were recorded and utilized in this study. The Duke operating rooms utilized 2 channel unilateral BIS electrodes that were connected via an E-BIS module to display the BIS index and raw waveform on the anesthesia GE (General Electric) monitors. The Mt. Sinai cases utilized BIS Vista monitors (Covidien, Mansfield, MA).

At both institutions, the BIS proprietary algorithm transformed raw EEG data to a number from 0-100, with >90 indicating an awake state, <60 indicating unconsciousness and general anesthesia, <40 indicating deep sedation, and 0 indicating electrical silence or “burst suppression”. BIS values were obtained in a similar fashion from 5 minutes after incision until 5 minutes before the “end of surgery” time stamp, again to target the anesthetic “plateau phase”. The BIS index was recorded once per minute, and the median value from each 5-minute case epoch was used to calculate a case average mean, as described previously.^14^

### Inhaled Anesthetic Resistance Measurement

To gauge the appropriateness of BIS index values for a given aaMAC dose, and to measure the degree of BIS index drop for a given aaMAC dose, we developed the Duke Anesthesia Resistance Scale (DARS), defined as:

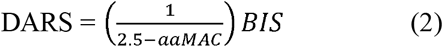

Here, *BIS* = mean of the median BIS readings during the case. The constant term of 2.5 represents the highest aaMAC value given in over 17,000 cases performed at our institution over a roughly two year period,^14^ and approximates the highest aaMAC value used in typical adult anesthesiology practice. A high DARS value could thus result from a high BIS reading and/or a large aaMAC. Conversely, a lower DARS value could result from a lower BIS reading and/or a small aaMAC.

### Delirium Evaluation and Diagnosis

All Duke patients in this study were followed daily after their index surgery by fellowship-trained geriatricians,^17^ all of whom underwent detailed training during their geriatrics fellowships on the standard DSM-V clinical criteria for making the diagnosis of delirium. These attending geriatricians closely examined patients for delirium based on the Confusion Assessment Method and then coded delirium (if present) with one of the following ICD9 codes in the patient’s chart: 290.11, 290.3, 290.41, 291.0, 292.81, 293.0, 293.1, 298.2, 348.3, 348.31, 348.39, 349.82, 437.2, 572.2, 768.7, 768.71, 768.72, 768.73, 780.09, or 780.97.^13^ Duke derivation cohort patients were defined as having postoperative delirium if any of these ICD9 codes was present in their patient record at any point during their postoperative index hospitalization. No inter-rater delirium reliability assessments were conducted between attending geriatricians in the Duke cohort, as attending level geriatrician or psychiatrist assessments are already considered the “gold standard” for the evaluation of delirium.^20^

For Mt. Sinai patients, delirium assessments were performed twice daily by research study staff using the CAM-ICU instrument. Delirium assessment training for Mt. Sinai study staff was performed by the neuropsychiatry team; inter-rater reliability for Mt Sinai staff delirium assessments was not performed because the CAM-ICU questions have very little subjectivity.^21^ Mt. Sinai patients were considered to have postoperative delirium if they had a positive delirium assessment at any point during hospitalization after their index surgery.

At both Duke and Mt Sinai, in order to avoid potential bias in delirium assessments, individuals performing the delirium assessments were blinded (i.e. not given access) to intraoperative DARS scores.

### Anticholinergic cognitive burden (ACB) score

The ACB assigns prescribed medications with known anticholinergic activity a score from 0-3 based on degree of predicted cognitive impairment in older adults, based on a multi-disciplinary consensus opinion validated to predict adverse outcomes.^22,23^

### Statistical analysis

Initial analysis was conducted just on the derivation cohort (i.e. Duke POSH patients). Categorical and numeric patient characteristics were summarized and compared between patients testing positive and negative for delirium with Pearson Chi-Square, Fisher Exact, t-tests or Wilcoxon rank sum tests as appropriate. Normality was assessed via Shapiro-Wilks tests and non-parametric statistics were used when evidence of non-normality was found. We examined the association of numeric DARS with delirium status as well as the pattern of empirical log-odds for delirium by DARS quintile.

We found evidence of a threshold effect in the empirical log-odds results; thus, we used ROC curve analysis and the Youden Index^24^ in order to identify the optimal cut point for DARS associated with delirium incidence in the derivation cohort. The Youden index captures the performance of the DARS as a function of sensitivity and specificity for classification of a binary outcome. The Youden index (*J*) is defined as:

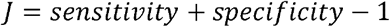

The optimal cut point is the DARS value that maximizes the value of *J*. We determined the association of the created binary DARS variable via chi-square test and odds ratio estimation. Subsequently we performed univariable and multivariable logistic regression for delirium outcome based on the binary DARS variable adjusting for potential confounding from *a priori* known delirium risk factors such as age, procedure duration, and anticholinergic burden score. Using empirical logits we explored the functional relationship between the numeric factors and delirium and found only DARS to have a significant non-linear relationship. Since our observed delirium incidence was low, we used Firths penalized likelihood in our multivariable logistic regression analysis to control for multiple potential confounding factors.

Following analysis and identification of the optimal DARS cut point for delirium prediction in the derivation cohort, we measured the incidence of a DARS <27 in the Mt. Sinai cohort and repeated the univariable and multivariable analysis of its association with postoperative delirium. Given the limited sample size available from each institution, we also conducted an analysis of the combined cohort to obtain more stable and generalizable effect estimates. For the combined cohort analysis we employed generalized mixed model techniques, including a random effect for institution, to repeat the univariable and multivariable analysis of the association of DARS <27 with postoperative delirium. To evaluate the diagnostic and predictive performance of DARS < 27 we report sensitivity, specificity, positive predictive value and negative predictive value.

This was an exploratory study designed to develop a composite processed EEG-based measure of brain resistance to inhaled anesthetics and evaluate the relationship between such a measure and postoperative delirium. At the time the project was conceived, there was no prior literature describing either a processed EEG measure of brain anesthetic resistance, nor describing the relationship between such a measure and postoperative delirium risk. Thus, there was insufficient data to perform a detailed *a priori* power analysis for this exploratory study, Nonetheless, we reasoned that this study would likely have sufficient power, as prior studies relating other EEG metrics to other neurocognitive variables have often had slightly smaller sample sizes^11,25^ than each of the individual cohorts studied here.

All statistical analyses were perfomed using SAS v 9.4 (SAS Inc., Cary, NC), and p < 0.05 was considered statistically significant.

## RESULTS

We identified 69 Duke patients followed by the preoperative optimization of senior health clinic (derivation cohort) and 70 Mt Sinai patients (validation cohort) who met our inclusion criteria (consort diagram Figure 1A-B). Baseline and intraoperative characteristics are shown in Table 1. Postoperative delirium occurred in 32% (n=22) of the Duke derivation cohort and 19% (n=13) of the Mount Sinai validation cohort (Table 1). The derivation cohort patients tended to have shorter procedure length (mean=139 mins) than validation cohort patients (mean=162 mins). The case average aaMAC tended to be slightly lower for the derivation cohort patients (mean=0.85) versus the validation cohort patient (mean=1.01). The mean ACB score was zero in both cohorts.

**Table 1.** Duke Derivation cohort and Mount Sinai Validation cohort patient characteristics. Intraoperative medication data was not available from the validation cohort at Mt. Sinai. In the Duke cohort, 2 patients were missing Anti-Cholinergic Burden scores; no other data was missing. Opioid dosage is given in oral morphine equivalents, non-depolarizing paralytic dosage is given in rocuronium mg equivalents (see supplemental methods for details). Numeric variables are summarized with mean (SD) or median [Q1, Q3] and categorical variables with N (%). ASA, American Society of Anesthesiology. BIS, bispectral index. OME, oral morphine equivalents. N_2_O, nitrous oxide.

|  | Duke (N=69) | Mt Sinai (N=70) |
| --- | --- | --- |
| Age | 73 [69, 79] | 70 [67, 75] |
| Gender (Male) | 31 (44.9%) | 29 (41.4%) |
| Body Mass Index (kg/m<sup>2</sup>) | 27.6 [24.7, 31.6] | 27.3 [24.0, 33.9] |
| Weight (kg) | 76.3 [68.0, 88.5] | 81.6 [63.0, 97.5] |
| ASA Status | 3 [3, 3] | 3 [2, 3] |
| Anti-Cholinergic Burden Score | 0 [0, 1] | 0 [0, 1] |
| General surgery | 18 (26.1%) | 41 (58.6%) |
| Orthopedic surgery | 50 (72.5%) | 0 (0.0%) |
| Thoracic surgery | 0 (0.0%) | 16 (22.9%) |
| Urologic surgery | 1 (1.4%) | 13 (18.6%) |
| Procedure Length (min) | 139 [105, 194] | 162 [117, 227] |
| Desflurane Used | 18 (26.1%) | 24 (34.3%) |
| Isoflurane Used | 30 (43.5%) | 26 (37.1%) |
| Sevoflurane Used | 21 (30.4%) | 20 (28.6%) |
| Case Average age adjusted end tidal MAC fraction | 0.858 [0.746, 0.982] | 1.01 [0.901, 1.278] |
| Age adjusted end tidal MAC fraction Hours | 1.85 [1.21, 2.86] | 2.74 [1.92, 4.44] |
| Case Average BIS | 49.7 (10.2) | 47.0 (6.9) |
| Case Average BIS <45 | 25 (36.2%) | 25 (35.7%) |
| Propofol Used / Dose (mcg/kg/min) | 65 (94.2%) / 13.6 [9.6, 16.6] | -- |
| N<sub>2</sub>O used | 2 (2.9%) | -- |
| Non-depolarizing Paralytics Used /Dose (mg) | 66 (95.7%) / 60 [42, 80] | -- |
| Ketamine Used / Dose (mcg/kg/min) | 45 (65.2%) / 6.0 [4.8, 7.2] | -- |
| Opioids Used / Dose (ME mg) | 65 (94.2%) / 28 [20, 36] | -- |
| Phenylephrine Used / Dose (mg) | 62 (89.9%) / 1.8 [0.6, 4.5] | -- |
| Midazolam Used / Dose (mg) | 33 (47.8%) / 2 [1, 2] | -- |
| Epidural Used | 21 (30.4%) | -- |
| Dexmedetomidine Used / Dose (mcg) | 4 (5.8%) / 18 [12, 20] | -- |
| Mean Duke Anesthesia Resistance Scale Score | 30.61 [26.20, 35.64] | 34.01 [29.59, 42.22] |
| Duke Anesthesia Resistance Scale <27 | 20 (29.0%) | 13 (18.6%) |
| Delirium | 22 (31.9%) | 13 (18.6%) |

**Figure 1:**
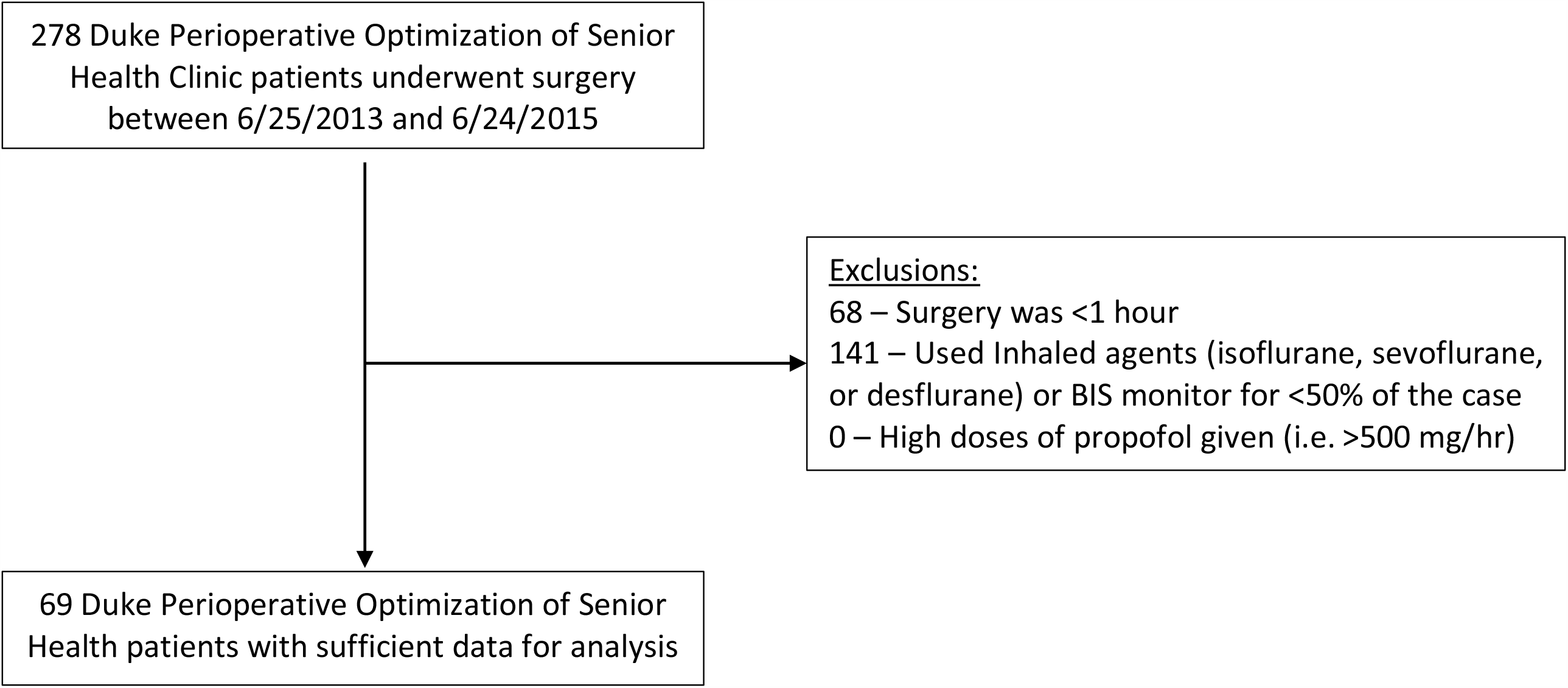
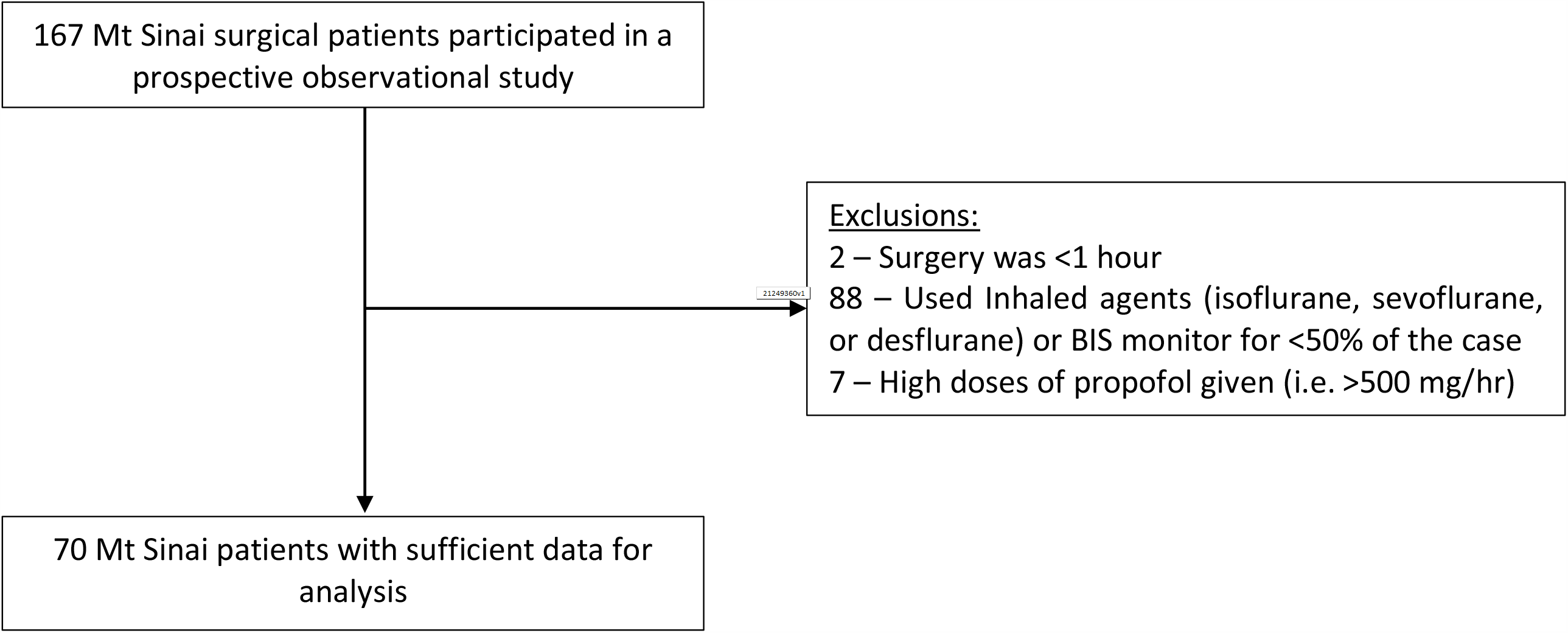
CONSORT Diagram for the Duke Derivation cohort (A) and the Mt Sinai Replication Cohort (B).

**Figure 2:**
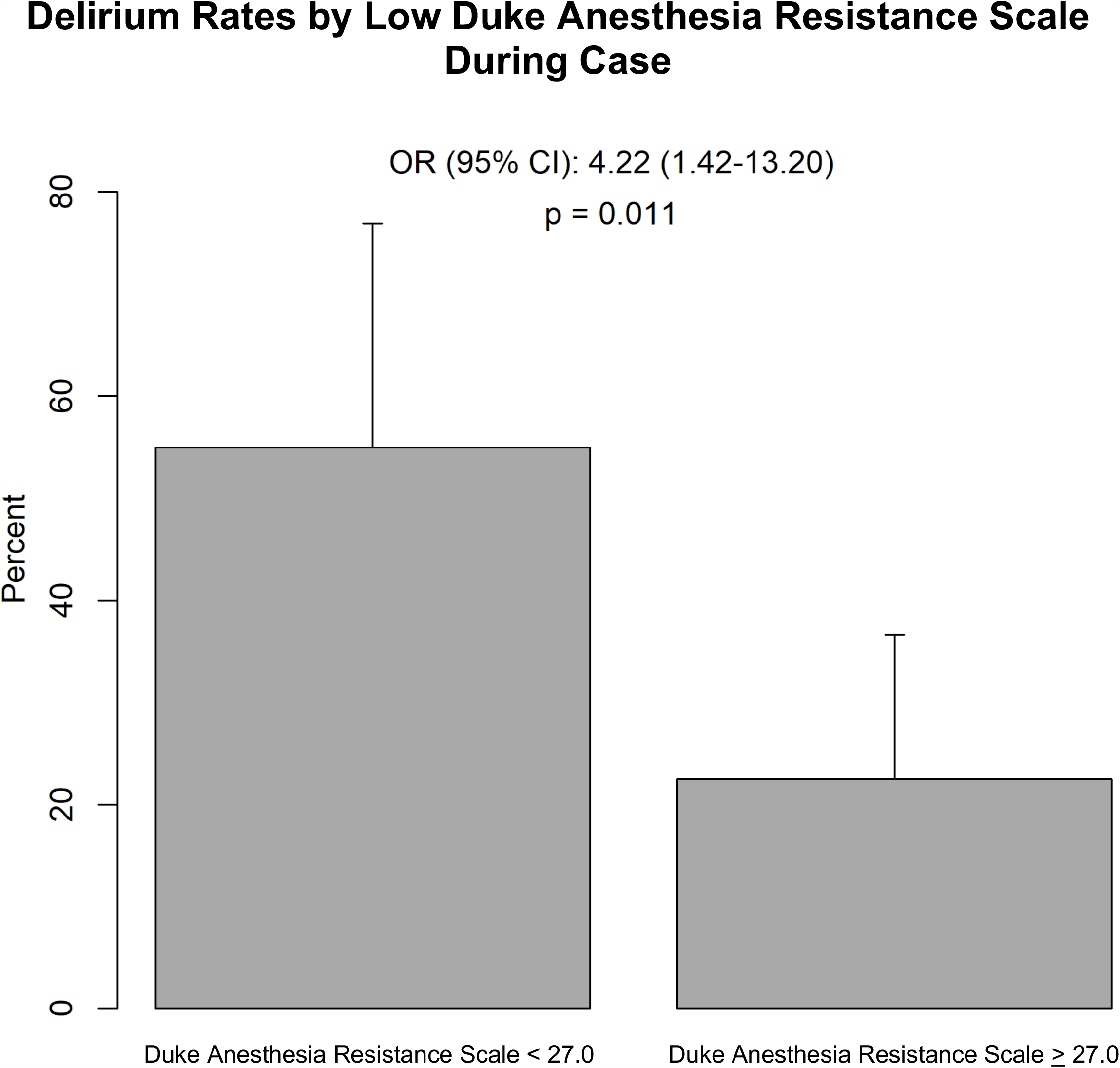
Increased Postoperative Delirium Rates in Patients with a DARS <27 in the Duke patient cohort (derivation cohort). Error bars represent the 95% confidence interval.

There were no statistically significant differences between patients with versus without delirium in the Duke derivation cohort patients (Supplemental Table 1), though patients who developed delirium tended to have longer surgeries. Age and ASA class were not significantly associated with delirium in the Duke POSH patients (derivation cohort). Neither aaMAC (median 0.9 vs. 0.8; p=0.593) nor case average BIS values (mean 51 vs. 48; p=0.300) differed among patients with vs without postoperative delirium. DARS scores were also not significantly different between patients with vs without delirium (median of 27.3 vs. 31.9, p=0.075; Supplemental Table 1). Based on the empirical logits we found that the relationship between DARS score and postoperative delirium was highly non-linear in Duke patients (derivation cohort; Supplemental Table 2).

To determine an optimal test threshold for the DARS value, an ROC curve was created (Supplemental Figure 1) and the Youden index was calculated at various cutoffs. A DARS value of 27.0 was found to optimize the Youden index and had high specificity for postoperative delirium (0.81). This binary DARS cutoff yielded a positive predictive value of 0.55 and a negative predictive value of 0.78 for postoperative delirium in the Duke derivation cohort.

Univariate logistic regression models of DARS <27.0 and postoperative delirium for the derivation and validation cohorts are shown in Table 2. The incidence of a DARS < 27 in the derivation cohort was 29% (n=20) and 19% (n=13) in the validation cohort. In the derivation cohort, patients with a DARS <27.0 had a four-fold increased odds of developing postoperative delirium (OR 4.2, 95% CI: 1.42 – 13.2; p=0.011). In the validation cohort, patients with a DARS <27.0 had a greater than six-fold increased odds of developing postoperative delirium (OR 6.1, 95% CI: 1.59 – 24.4; p=0.008). For the combined cohorts, a DARS < 27.0 was associated with a five-fold increase in the odds of postoperative delirium (OR 5.0, 95% CI: 2.12 – 11.9; p<0.001).

**Table 2:**
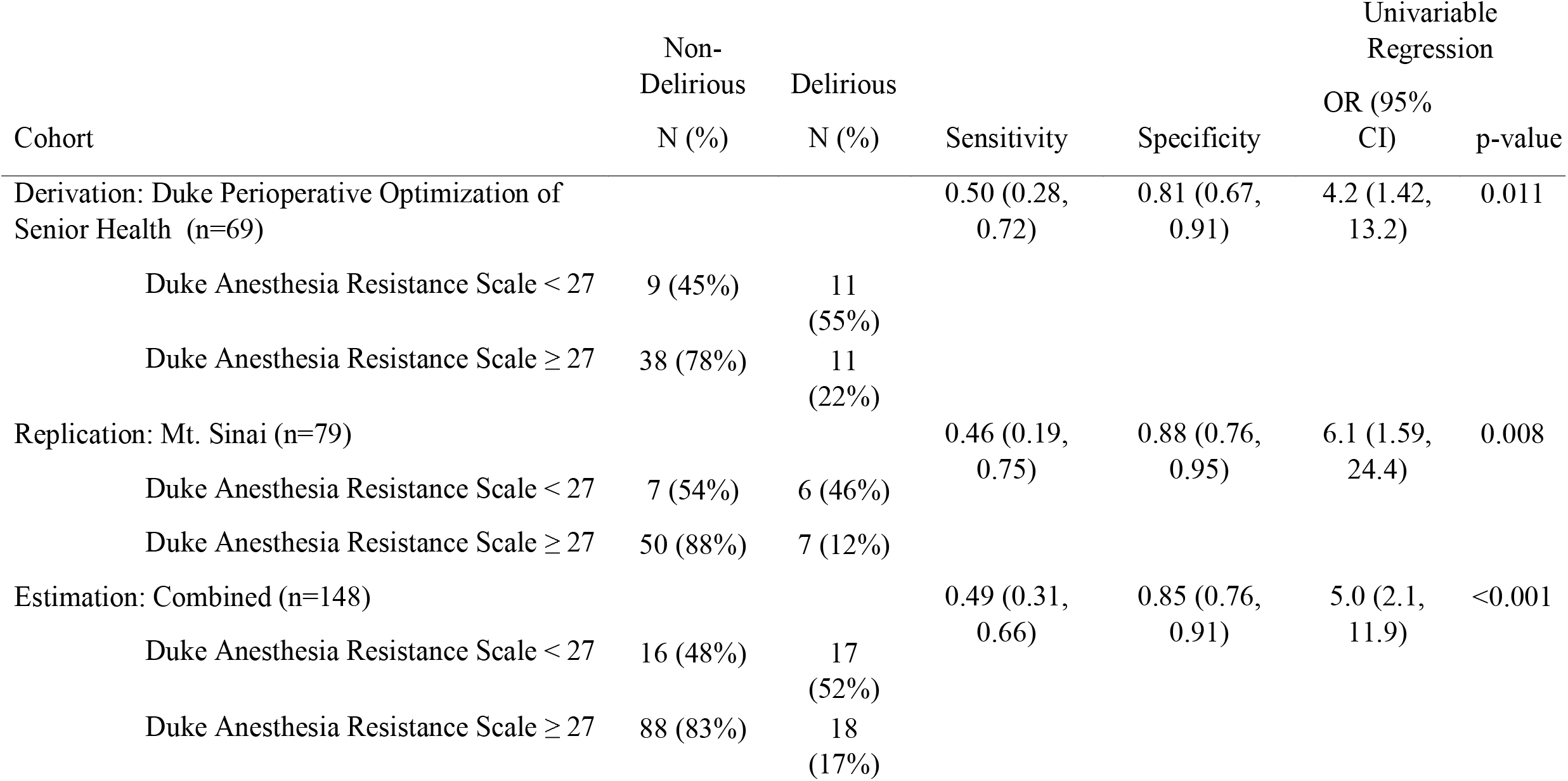
Univariable relationship between Duke Anesthesia Resistance Scale (<27.0) and Postoperative Delirium in the derivation, validation, and combined estimation cohorts. Logistic regression was used for the derivation and validation cohorts, and a generalized linear mixed model was used for the combined estimation cohort with a random intercept term for site. Numbers in parentheses represent 95% confidence intervals. Validation

In multiple variable regression models adjusting for preoperative and intraoperative factors previously shown to be associated with postoperative delirium (Table 3), a DARS score <27.0 remained associated with an increase in the odds of postoperative delirium in each of the cohorts (combined estimation cohort OR 4.7, 95% CI: 1.87–12.0; p<0.001). In the combined estimation cohort, for each additional ten minutes of anesthetic/surgical duration the odds of postoperative delirium increased by a factor of 1.07 (95% CI 1.01 – 1.14; p=0.028). To provide additional estimates of effects and uncertainties in the derivation cohort, we also examined the multivariable analysis in 1000 bootstrap replicates; this demonstrated a median odds ratio and empirical 95% confidence interval for a DARS < 27 predicting delirium of 4.8 [1.03, 29.6].

**Table 3:** Multivariable Logistic Regression Prediction Model for Postoperative Delirium (with Firth Correction due to low event rate) in the derivation and validation cohorts, and from a generalized linear mixed model for the combined cohort (random intercept for site). ASA, American society of Anesthesiology.

| Effect | Derivation: Duke<br>Perioperative Optimization<br>of Senior Health |  | Validation: Mt. Sinai |  | Estimation: Combined |  |
| --- | --- | --- | --- | --- | --- | --- |
|  | OR (95% CI) | P-Value | OR (95% CI) | P-Value | OR (95% CI) | P-Value |
| Duke Anesthesia<br>Resistance Scale < 27 | 3.53 (1.08, 12.2) | 0.047 | 5.1 (1.33, 20.7) | 0.025 | 4.7 (1.87, 12.0) | 0.001 |
| Age (per year) | 1.05 (0.96, 1.14) | 0.295 | 1.04 (0.93, 1.16) | 0.469 | 1.05 (0.98, 1.12) | 0.184 |
| ASA Status (per unit) | 1.21 (0.26, 5.5) | 0.814 | 0.57 (0.15, 2.07) | 0.394 | 0.76 (0.27, 2.09) | 0.588 |
| Male vs. Female Sex | 0.74 (0.21, 2.49) | 0.633 | 0.95 (0.26, 3.32) | 0.940 | 0.80 (0.32, 1.97) | 0.616 |
| Procedure Duration (per<br>10 min) | 1.09 (1.01, 1.20) | 0.047 | 1.05 (0.96, 1.15) | 0.310 | 1.07 (1.01, 1.14) | 0.028 |
| Anticholinergic Burden<br>Score | 1.46 (1.05, 2.21) | 0.051 | 0.97 (0.47, 1.68) | 0.914 | 1.33 (0.99, 1.78) | 0.060 |

A DARS value <27.0 also remained a significant independent predictor of delirium risk in the derivation cohort (OR 5.4, 95% CI: (1.6, 21.1); p<0.014) while accounting for dosage of other anesthetic drugs that can affect EEG parameters (i.e. opioids,^26^ non-depolarizing neuromuscular blockers,^27^ ketamine,^28^ midazolam and propofol^29^) and the use of epidural anesthesia^30^ and/or nitrous oxide^31^ (Table 4). None of these other variables were significantly associated with postoperative delirium risk.

**Table 4:**
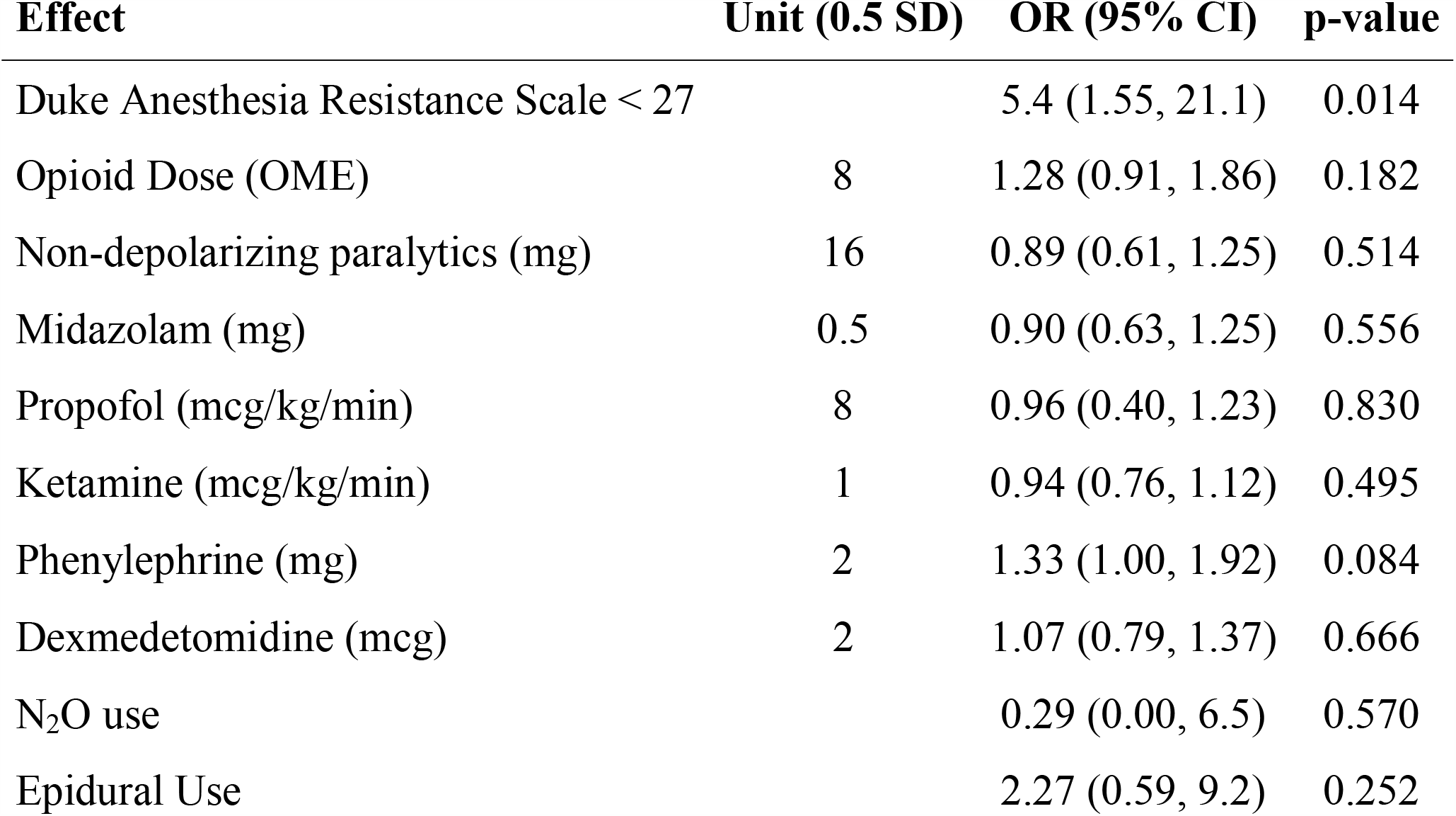
Multivariable Logistic Regression Model for Postoperative Delirium (with Firth Correction due to low event rate) in the derivation cohort accounting for intraoperative drugs. Units represent 0.5 SD of the mean of each variable, rounded to 1 significant figure. Non-depolarizing paralytics are measured in rocuronium equivalent mg (see supplemental methods for details). OME, oral morphine equivalents. N_2_O, nitrous oxide.

## DISCUSSION

In this dual center study in older adults, we found that a processed EEG measure of decreased brain resistance to anesthetic dose-related reductions in neurophysiologic activity (i.e. the DARS) was an independent and reproducible predictor of postoperative delirium. As compared to patients with intraoperative DARS scores ≥27, those with a DARS score <27.0 in the combined cohort had over five times higher odds of developing postoperative delirium. Interestingly, even though neither BIS nor aaMAC alone was predictive of postoperative delirium risk, the DARS (a combined index based on both BIS and aaMAC) was highly predictive of postoperative delirium. The DARS effectively scales the BIS score (in which lower scores indicate a less active brain state) by the difference between the maximum aaMAC fraction likely used in clinical practice minus the actual aaMAC fraction received by the patient. A lower DARS score is thus indicative of a less active brain state (i.e. a lower BIS score) than would typically be expected for a given inhaled anesthetic dose (i.e.aaMAC fraction), i.e. decreased brain resistance to anesthetic-induced decreases in brain activity.

Roughly 50% of patients with a DARS score < 27.0 had delirium in both the derivation and validation cohorts, while only 12-22% of patients in each cohort with a DARS score >27.0 had delirium. In both cohorts, a DARS score < 27 was roughly 50% sensitive but more than 80% specific for predicting delirium. Thus, a DARS score < 27.0 can be used clinically to direct scarce resources (such as psychiatry or geriatrics consultations) towards patients at relatively higher risk for developing postoperative delirium.

The ∼50% sensitivity of the DARS for delirium risk prediction fits with the view that delirium results from a mixture of predisposing and precipitating factors.^32^ We hypothesize that lower brain anesthetic resistance (i.e. DARS <27.0) is an intrinsic characteristic of dysfunction within some patients’ brains (i.e. an intraoperative marker of preoperative brain dysfunction), and thus a predisposing factor for delirium. However, patients who develop delirium predominantly due to post-operative precipitating factors (e.g. pain, sleep disruption, etc) would not be expected to have a DARS < 27. Thus, it is unsurprising that a DARS < 27 has only 50% sensitivity for delirium prediction; a DARS < 27 likely only predicts delirium cases driven largely by predisposing factor(s) (i.e. the neuro-pathophysiologic factors that give rise to lower brain anesthetic resistance).

The DARS was a significant predictor of delirium regardless of which type of BIS monitor was used (i.e. E-BIS module displaying BIS values on GE anesthesia machines, vs BIS Vista), and independent of other anesthetic adjunct drugs used (Table 4). This robustness of the DARS for predicting delirium, despite varying doses of other drugs that can affect the EEG (i.e. opioids,^26,33^ ketamine,^28^ non-depolarizing neuromuscular blockers,^27^ nitrous oxide,^31^ etc), suggests that lower brain anesthetic resistance in response to volatile anesthetics is relatively greater in magnitude than the EEG changes due to these other anesthetic drug adjuncts. Further, the DARS was an independent predictor of delirium in two separate cohorts from two separate institutions, in whom delirium was assessed by two different groups of personnel (geriatricians vs trained research staff) using two different instruments (regular CAM vs CAM-ICU). Despite all of these differences, the DARS was a robust predictor of delirium in both cohorts, suggesting that it identifies core neurologic features of a brain at risk for postoperative delirium independent of specific intraoperative anesthetic practices, BIS monitor types, or particular delirium assessment tools.

The data presented here complement the recent finding that increased anesthetic sensitivity, as measured by burst suppression divided by a composite measure of anesthetic dosage, predicts postoperative delirium risk.^13^ Indeed, BIS values <30 are linearly (and inversely) related to burst suppression ratio.^34^ Thus, the relationship identified here between lower DARS scores (while are likely due in part to lower BIS scores) and increased postoperative delirium risk may in part reflect an association between increased burst suppression at lower inhaled anesthetic doses and increased postoperative delirium risk, as Fritz and colleagues reported.^13^ Nonetheless, the data presented here simply show an association between DARS and delirium; these data do not demonstrate that a DARS < 27 “causes” delirium. Indeed, the fundamental neurobiological etiology of delirium remains to be elucidated.

In this study, lower brain anesthetic resistance (i.e. a DARS score) < 27 was a better predictor of delirium risk than chronologic age. This finding is similar to the recent finding that neurophysiologic brain age is a better predictor of delirium risk than chronologic age.^7^ Both of these findings are consistent with the general principle of geriatric medicine that the variance in the function of most organs across the population increases with age,^35^ and variability in organ function within an individual patient cannot be entirely predicted by chronological age alone. Decreased brain resistance to anesthesia, which can also be viewed reciprocally as increased brain sensitivity to anesthesia, may relate to the concept of increased “brain age” in some patients. Indeed, there are age-dependent changes in EEG responses to inhaled anesthetics as well as propofol,^36^ especially in older adults.^37^ Pivotal work by Purdon and colleagues^37^ focused on chronological age-dependent changes in EEG responses to anesthesia, yet chronological age can clearly be disassociated from biological age,^38,39^ especially for the brain.^40^ For example, anesthetic-induced frontal alpha power decreases with age,^37^ yet there is still substantial variance in alpha power among older adults.^8^ In fact, Hesse and colleagues found that markers of “brain age” can predict postoperative delirium better than chronological age, perhaps due to age-dependent biological changes that occur within the brain.^7^ Interestingly, chronological age was not a significant predictor of delirium in this study, which likely reflects the relative narrow age distribution of the cohorts studied here (i.e. virtually all patients in both cohorts were in their late 60’s or older).

Biological age is related to changes in processes such as DNA stability, protein modifications, immunologic activity, metabolic and oxidative stress, and increased inflammation,^39^ which has been referred to as “inflammaging”.^41^ In fact, inflammation increases anesthetic sensitivity in both cultured neurons and whole animals.^42^ Thus, increased brain inflammation at baseline and/or in response to surgical stress^43,44^ could make the brain less resistant to anesthesia, which would result in lower DARS values. Our results and recent findings^13^ suggest that lower brain resistance to anesthesia is associated with postoperative delirium risk. Since delirium occurs most often in older adults, an important question for future study is to what extent inflammation within the brain increases with age (either overall or specifically in some patients) versus to what extent the aging brain is less resistant to similar levels of neuro-inflammation as a younger brain. Either of these two conceptual possibilities could help explain a potential link between brain inflammation, age, and postoperative delirium risk.

Aside from these two possibilities, this study has three limitations. First, the patient cohorts from both institutions are of moderate size. To address this limitation, we analyzed the cohorts both separately and jointly, and employed Firth’s penalization in our regression models. The separate analyses demonstrates the utility and reproducibility of a DARS < 27 for predicting delirium risk in each of the two cohorts, while the joint analysis maximizes sample size to provide a more robust and generalizable effect size estimate. We also pursued bootstrapping to improve estimates of effects and uncertainties, however, given the low number of delirium events and incidence of DARS < 27 in our derivation cohort, the estimates were no more precise than in our primary analysis. Second, as mentioned above, the two cohorts studied here utilized different methods for identifying postoperative delirium. Delirium was identified by geriatrician interview (based on DSM-V criteria) in the Duke cohort, while delirium was identified by CAM assessments performed by research staff in the Mt. Sinai cohort. Nonetheless, the use of these two different delirium assessment methods in these two cohorts is a strength of this study, because it suggests that a DARS score < 27 predicts increased delirium risk independent of the specific delirium assessment method used. Nonetheless, further study is necessary to validate the ability of lower DARS to predict increased delirium risk in larger and more heterogeneous cohorts.

Third, the DARS utilizes the BIS index, a proprietary processed index with an underlying algorithm that has never been published, and which has not been validated for its original intended purpose of reducing intraoperative awareness.^15,16^ Further, we have recently shown that BIS values may be erroneously high in older adults.^14^ Nonetheless, BIS monitors are used in 2.7 million surgical cases per year (V. Curro of Covidien/Medtronic, personal communication, Nov 8, 2018). In the current absence of an “ideal” intraoperative anesthetic EEG monitor,^4,45^ and despite these shortcomings of the BIS, the DARS offers a way to utilize available BIS data together with end tidal anesthetic dosage to predict postoperative delirium risk in older patients. Nonetheless, larger studies at multiple centers are necessary to study further this relationship between brain anesthetic resistance and delirium risk. Additionally, further studies are warranted to determine if other EEG parameters (such as alpha power, entropy measures, etc) would have similar or greater utility than the processed BIS index for using in brain anesthetic resistance equations to predict postoperative delirium.

The DARS represents an opportunity to identify patients at higher risk for delirium and modify perioperative care accordingly. Delirium prevent strategies can be time-consuming and resource-intensive, so they are generally reserved for patients that are flagged as “high delirium risk” by preoperative screening programs.^17,46,47^ The DARS could allow anesthesiologists to identify patients in the operating room who are at increased risk for developing postoperative delirium. For example, consider two 70 patients both receiving 1.6% sevoflurane (∼1 aaMAC) who have average intraoperative BIS values of 30 versus 50. The patient with a BIS of 30 would have a DARS score of 20 [30/(2.5-1)=20], while the patient with a BIS of 50 would have a DARS score of 33.3 (50/(2.5-1)=33.3]. Based on our combined estimation cohort results (Table 2), the patient with a DARS of 20 would have a 52% chance of developing delirium, while the patient with a DARS of 33 would have a delirium risk of only 17%. Calculating the DARS could thus allow institutions to allocate resources to the most at-risk patients by optimizing postoperative inpatient delirium prevention, medication management, and anticipation of home care needs.

In summary, we have developed a processed EEG-based measure of brain anesthetic resistance that predicts postoperative delirium risk. Further investigation is necessary to replicate these findings, to understand better their neurophysiologic/mechanistic basis, to determine the relationship between DARS score and other postoperative adverse outcomes, and to study whether altering intraoperative care in patients with lower DARS scores could help prevent delirium in this high-risk population.

## Data Availability

The authors confirm that the data supporting the findings of this study are available within the article and its supplementary materials.

## Acknowledgements

We thank Austin Traylor (Duke Anesthesiology Department) for obtaining intraoperative electronic data from the Epic medical record system, and Dr Stacey Chung (Duke Anesthesiology Department) for literature search assistance. The Duke Anesthesia Resistance Scale, including the equation, and all related documentation and software, are owned by Duke University. (c) Copyright 2020. Duke University. All Rights Reserved. Developed by Duke University School of Medicine, Duke University.

