## Supplemental Figure 1 for "A Processed EEG based Brain Anesthetic Resistance Index Predicts Postoperative Delirium in Older Adults: A Dual Center Study"

**Supplemental Figure 1:** ROC Analysis for Duke Anesthesia Resistance Scale and Postoperative Delirium in the Duke Perioperative Optimization of Senior Health cohort. The area under the curve (AUC) is estimated as 0.63 (0.48-0.79) p=0.08. Youden index indicates optimal cut point is at Duke Anesthesia Resistance Scale value of 27.0, thus defining a low Duke Anesthesia Resistance Scale as < 27.0.

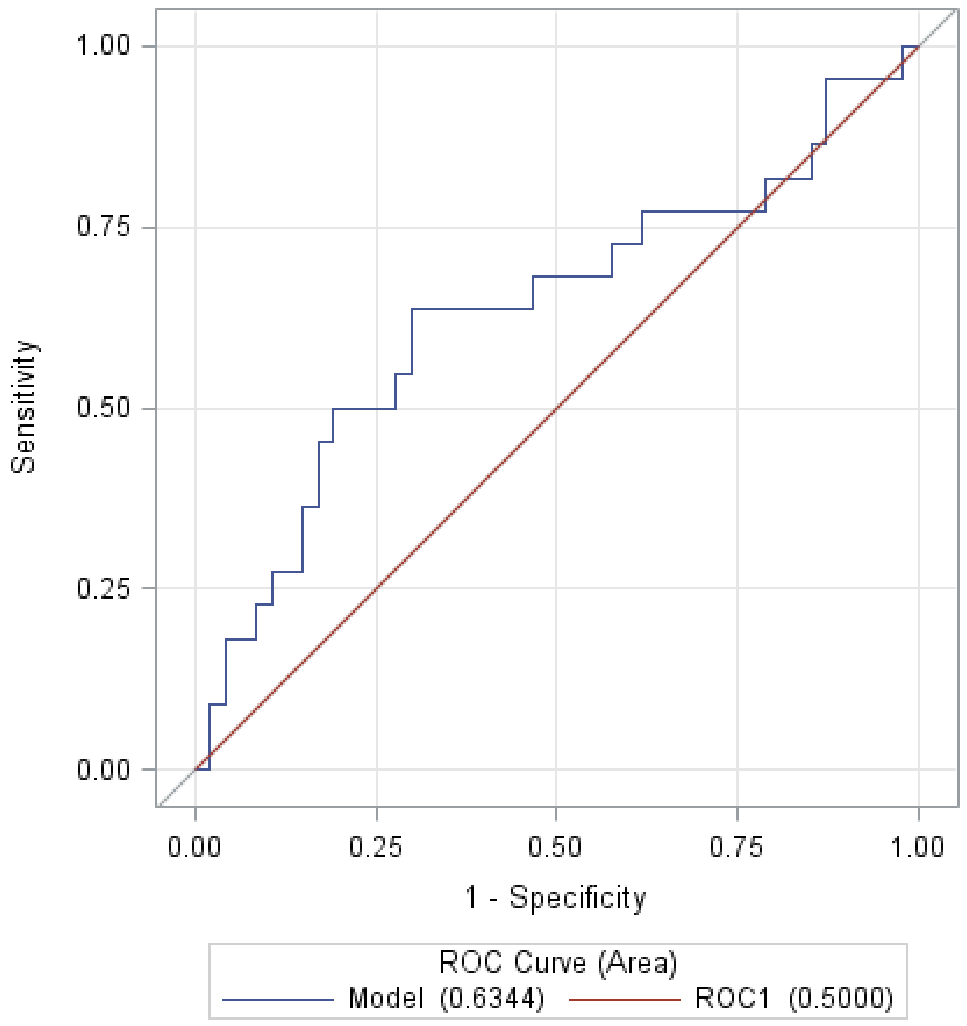
