## Supplemental Table 1 for "A Processed EEG based Brain Anesthetic Resistance Index Predicts Postoperative Delirium in Older Adults: A Dual Center Study"

**Supplemental Table 1:** Baseline and Intraoperative Patient Characteristics for the Derivation Cohort (Duke Perioperative Optimization of Senior Health patients), separated by delirium status.

|  | Delirium Negative  (N=47, 68%) | Delirium Positive  (N=22, 32%) | Total  (N=69) | p value |
| --- | --- | --- | --- | --- |
| Age | 72 (69, 79) | 74 (70, 82) | 73 (69, 79) | 0.3699^1^ |
| Gender (Male) | 22 (46.8%) | 9 (40.9%) | 31 (44.9%) | 0.6462^2^ |
| Body Mass Index | 27.8 (24.3, 31.6) | 27.2 (24.7, 32.1) | 27.6 (24.7, 31.6) | 0.8874^1^ |
| Weight (kg) | 74.4 (67.3, 92.1) | 80.2 (73.5, 88.0) | 76.3 (68.0, 88.5) | 0.5113^1^ |
| ASA Class |  |  |  | 0.1855^2^ |
| 2 | 6 (12.8%) | 1 (4.5%) | 7 (10.1%) |  |
| 3 | 37 (78.7%) | 21 (95.5%) | 58 (84.1%) |  |
| 4 | 4 (8.5%) | 0 (0.0%) | 4 (5.8%) |  |
| Total # of baseline Medications | 12 (8, 15) | 10 (8, 14) | 11 (8, 15) | 0.5437^1^ |
| Anticholinergic Burden Score | 0 (0, 1) | 1 (0, 3) | 0 (0, 1) | 0.1574^1^ |
| Saint Louis University Mental Status Score^*^ | 24 (21, 27) | 22.5 (19, 28) | 24 (20, 27) | 0.3859^1^ |
| Baseline Cognitive Impairment^**^ | 19 (41.3%) | 10 (47.6%) | 29 (43.3%) | 0.6284^2^ |
| Prior History of Delirium*** | 11 (23.9%) | 4 (19.0%) | 15 (22.4%) | 0.6576^2^ |
| Surgery Type |  |  |  | 0.7094^2^ |
| Cardiovascular | 1 (2.1%) | 0 (0.0%) | 1 (1.4%) |  |
| Digestive | 12 (25.5%) | 4 (18.2%) | 16 (23.2%) |  |
| Integumentary | 1 (2.1%) | 0 (0.0%) | 1 (1.4%) |  |
| Musculoskeletal | 32 (68.1%) | 18 (81.8%) | 50 (72.5%) |  |
| Urinary | 1 (2.1%) | 0 (0.0%) | 1 (1.4%) |  |
| Procedure Length (min) | 128 (98, 171) | 185.5 (109, 225) | 139 (105, 194) | 0.0880^1^ |
| Case Average MAP | 84.1 (8.2) | 82.1 (6.6) | 83.5 (7.7) | 0.2995^3^ |
| Gas Used |  |  |  | 0.0994^2^ |
| Desflurane | 10 (21.3%) | 8 (36.4%) | 18 (26.1%) |  |
| Isoflurane | 19 (40.4%) | 11 (50.0%) | 30 (43.5%) |  |
| Sevoflurane | 18 (38.3%) | 3 (13.6%) | 21 (30.4%) |  |
| Case Average age adjusted end tidal MAC fraction | 0.869 (0.721, 1.043) | 0.848 (0.761, 0.902) | 0.858 (0.746, 0.982) | 0.5931^1^ |
| Case Average BIS | 50.6 (10.5) | 47.8 (9.7) | 49.7 (10.2) | 0.2996^3^ |
| Duke Anesthesia Resistance Score (BIS/(2.5- age adjusted end tidal MAC fraction)) | 31.9 (27.4, 36.0) | 27.3 (24.9, 33.4) | 30.6 (26.2, 35.6) | 0.0745^1^ |
| Case Average BIS < 45 | 15 (31.9%) | 10 (45.5%) | 25 (36.2%) | 0.2755^2^ |
| *Missing for 10 patients, 8 of whom had sweet 16 administered instead  **Baseline Impairment defined as either a Saint Louis University Mental Status <24 or Sweet 16<14, missing for 2 patients  ***missing for 2 patients  ^1^Wilcoxon    ^2^Chi-Square    ^3^Equal Variance T-Test | | | | |

ASA: American Society of Anesthesiologists, MAP: Mean Arterial Pressure, BIS: Bispectral Index
