## Supplemental Table 2 for "A Processed EEG based Brain Anesthetic Resistance Index Predicts Postoperative Delirium in Older Adults: A Dual Center Study"

**Supplemental Table 2**: Empirical Log Odds for Delirium based on Quintiles of the Duke Anesthesia Resistance Score distribution in the Duke Perioperative Optimization of Senior Health cohort (derivation cohort).

| Quintile | N | Mean  Duke Anesthesia Resistance Score | Min  Duke Anesthesia Resistance Score | Max  Duke Anesthesia Resistance Score | N  Delirium | Log  Odds | Prob |
| --- | --- | --- | --- | --- | --- | --- | --- |
| 1 | 13 | 22.9 | 17.1 | 25.5 | 6 | -0.143 | 0.481 |
| 2 | 14 | 26.9 | 25. 7 | 28.5 | 7 | 0.000 | 0.517 |
| 3 | 14 | 30.5 | 28.8 | 32.5 | 2 | -1.609 | 0.172 |
| 4 | 14 | 34.4 | 32.6 | 36.7 | 3 | -1.190 | 0.241 |
| 5 | 14 | 42.2 | 37.9 | 53.4 | 4 | -0.847 | 0.310 |
