## Supplemental Methods for "A Processed EEG based Brain Anesthetic Resistance Index Predicts Postoperative Delirium in Older Adults: A Dual Center Study"

*Duke Perioperative Optimization of Senior Health Clinic***:** This clinic provides multidisciplinary preoperative assessment and postoperative geriatrician follow-up to improve outcomes for older adults undergoing elective surgery. The preoperative assessment team at this clinic includes a geriatrician, geriatric resource nurse, social worker, program administrator, and nurse practitioner from the Preoperative Anesthesia Testing clinic, and includes a comprehensive preoperative geriatric evaluation. The team actively engages patients and their families in preoperative risk assessment and modification, focusing on specific “care points” considered crucial for optimizing care: cognition, medications, comorbidities, mobility, functional status, nutrition, hydration, pain, and advanced care planning. To facilitate implementation of preoperative clinic recommendations, the inpatient geriatrics team follows these patients after surgery and assists with managing medications, chronic conditions, pain, and common complications including delirium.

*Intraoperative Pharmacology*

Since the two main intravenous medium and long acting opioids administered in both patient cohorts were fentanyl and hydromorphone, the intravenous doses of these two drugs were converted into oral morphine equivalents and combined to give a total dose of medium and long acting intraoperative opioid administered (in oral morphine equivalents).

The main three non-depolarizing paralytics received by patients in the derivation (Duke) cohort were rocuronium, vecuronium and atracurium. Vecuronium and atracurium doses were converted into equivalent potency rocuronium doses by multiplying vecurium dosage by 6 and and atracurium dosage by 5/6^th^*,* thus allowing total non-depolarizing paralytic dosage (in rocuronium equivalents) to be used as a single term for analysis purposes.
